# A Comparative Analysis of Clinical Stage 3 COVID-19 Vaccines using Knowledge Representation

**DOI:** 10.1101/2021.03.07.21253082

**Authors:** Javier Burgos-Salcedo

## Abstract

The emergence of a novel SARS-CoV-2 coronavirus at the end of 2019 and its accelerated spread worldwide to become a pandemic has had, from the medical biotechnology point of view, an unprecedented global response, to the point that there are currently 176 vaccine candidates in preclinical stage, 66 in clinical stage, of which 19 are in phase 3, and 5 of these are massively applied worldwide. The purpose of the present work is to elaborate a hierarchical landscape of the current status of phase 3 vaccines, taking into account their attributes of technological platform, safety and efficacy. The methodology used was that of conceptual knowledge representation, resulting in, firstly, an appropriate classification of stage 3 vaccines, The Conceptual Lattice for COVID-19 vaccines, constructed according to how they relate to each other with respect to the set of their attributes. Secondly, the approach used allows proposing rational strategies for the design of heterologous vaccination schemes, which are urgently needed to control the pandemic.

## 1. Introduction

The severe acute respiratory syndrome coronavirus 2 (SARS-CoV-2) pandemic continues to spread throughout the world with explosive outbreaks underway throughout much of Europe and the United States [1]. In order to combat the disease, governments, international organizations, private companies, and academic institutions across the planet have launched a multifaceted vaccine development response with unprecedented rapidity. According with WHO [2,3], 176 vaccine candidates are in preclinical testing and at least 66 vaccines have already begun early clinical testing, and actually at least 19 vaccines are in phase 3 clinical development, and 10 vaccines are authorized to be used worldwide [4].

These facts indicate that effective vaccines have been emphasized to become the most effective strategy in the global fight against the COVID-19 pandemic [5,6], and given the actual number of promising COVID-19 vaccine candidates, elaborated from diverse technological platforms, with a broad range of safety and immunogenicity results reported in those individuals who underwent clinical trials, it is very important to build a rational landscape of phase 3 COVID-19 vaccines [7] focused towards an appropriate classification of them, leading to a functional hierarchy of these vaccines, from which concrete measures can be proposed for the optimization of dosage schedules, the minimization of reactogenicity and the increase of immunogenicity.

The aim of this study is to perform a comparative analysis of clinical stage 3 COVID-19 vaccines building a hierarchical model of them, based on its attributes related with safety, immunogenicity, dosing schedule and technological platform, using the theory of ordered sets and lattices, which are mathematical tools providing a natural setting in which to discuss and analyze such hierarchies. To meet the proposed objective, the Formal Concept Analysis (FCA) is used [8]; this is a method for knowledge representation and information management that is widely known among information scientists all around the world because its broad range of applications outside mathematics like economics, industry, chemistry, linguistics and environmental sciences among others [9-13].

Briefly, Knowledge Representation incorporates findings from psychology [14] about how humans solve problems and represent knowledge in order to design formalisms that will make complex systems easier to design and build, it is grounded on an understanding of human thinking based on concepts, which according to the main philosophical tradition, are constituted by its extension, comprising all the objects, in this case, a set of phase 3 COVID-19 vaccines, which belong to the concept, and its intension, including all attributes or properties v.gr. safety and immunogenicity, which applies to all objects of the extension.

The use of Knowledge Representation, through the Formal Concept Analysis, allows us to reach a series of conclusions about the vaccines for COVID-19 that are in phase 3 or already approved for use. In the first place, this set of vaccines can be represented as a hierarchical structure, in particular as a lattice, based on attributes such as technological platform, dosage, safety and immunogenicity. The conceptual analysis indicates that there is a direct relationship between the reactogenicity of the vaccines and their ability to induce neutralizing IgG antibodies: the higher the incidence of adverse effects, the higher the antibody titers are obtained. of course, reactogenicity generally consists of minor discomforts such as redness, fever, pain and others widely described elsewhere in the literature [15]. It should also be noted that the most effective dosage, regardless of the technological platform of the vaccine, is that of two doses delivered 21 days apart.

It should be noted that although all the vaccines considered in the present study have shown that they induce a protective humoral immune response against SARS-CoV-2 and are safe, clear differences can be observed in relation to the levels of neutralizing antibodies and the technological platform on which were made v.gr. DNA vaccines induce lower antibody titers in contrast to RNA vaccines, viral particles and viral-like particles, which in general are strongly reactogenic and produce high levels of neutralizing IgG. Non-replicating virus and inactivated virus vaccines induce medium to high antibody levels and exhibit less reactogenicity.

Finally, employing a principle of Formal concept Analysis, named duality, it was possible to establish the level of affinity within the set of COVID-19 stage 3 vaccines, making possible the development of a methodology for the elaboration of heterologous vaccination strategies for the control of the pandemic.

## 2. Materials and methods

### 2.1 COVID-19 phase 3 vaccines

The rapid development of vaccines to control COVID-19 has been closely followed by various organizations around the world, for February 12, 2021, the World Health Organization [] in its report Covid-19 Landscape of Novel Coronavirus Candidates Vaccines Development Worldwide lists a total of 176 vaccines in the preclinical stage, 66 in the clinical phase, of which 16 are in phase 3 clinical studies. Other entities such as the Milken Institute [16], report similar numbers, 248 vaccines in development, 56 are in the clinical phase, of which 10 are currently in use. On the other hand, various authors [5-7] report the technological, safety and immunogenicity characteristics of each of the phase 3 and / or authorized vaccines. Based on these reviews, **Table 1** is organized, with a total of 19 vaccines in phase 3 and / or authorized, to carry out the present study.

**Table 1.**
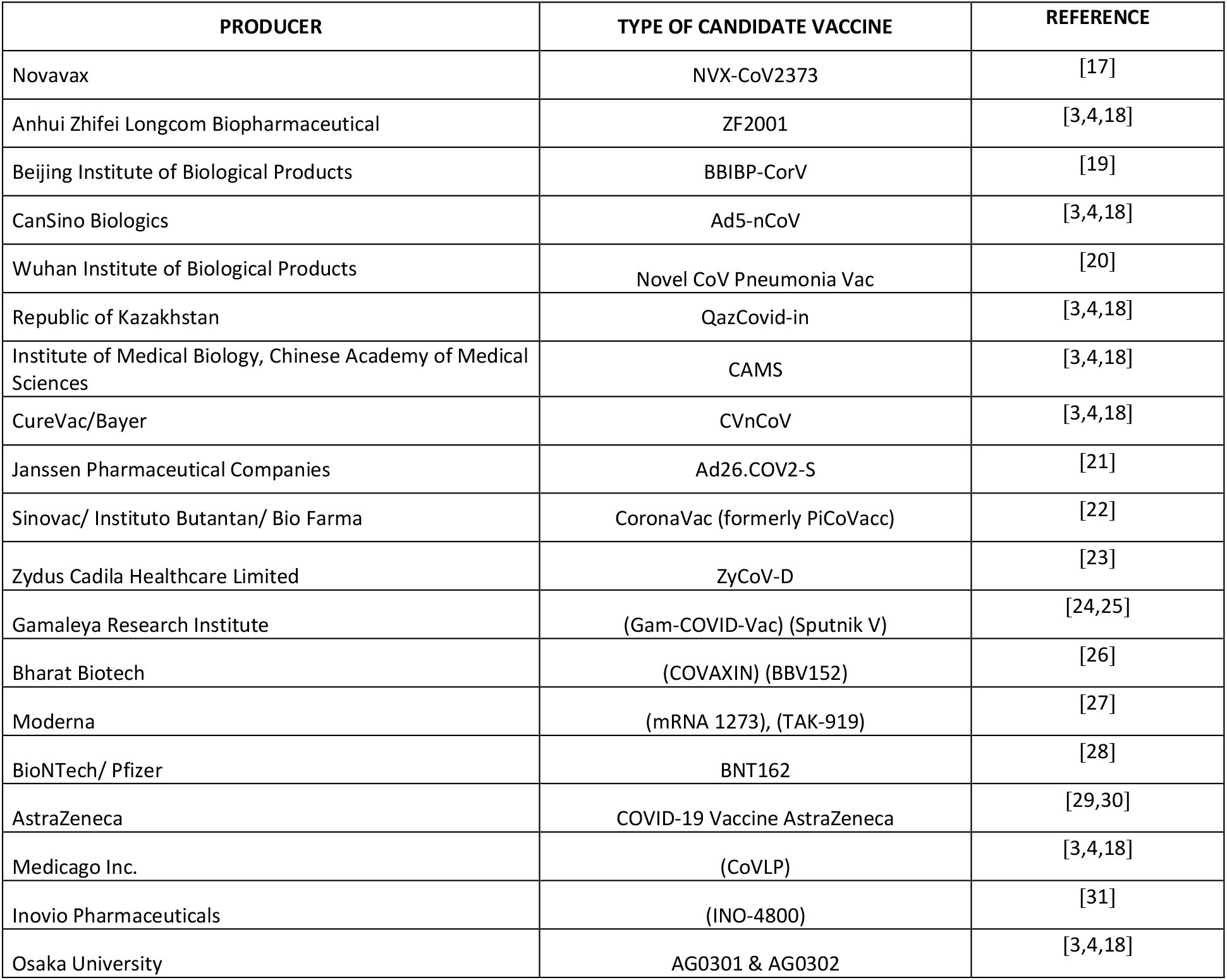
COVID-19 Vaccines in clinical phase 3 stage or authorized used for the present study.

**Table 2.**
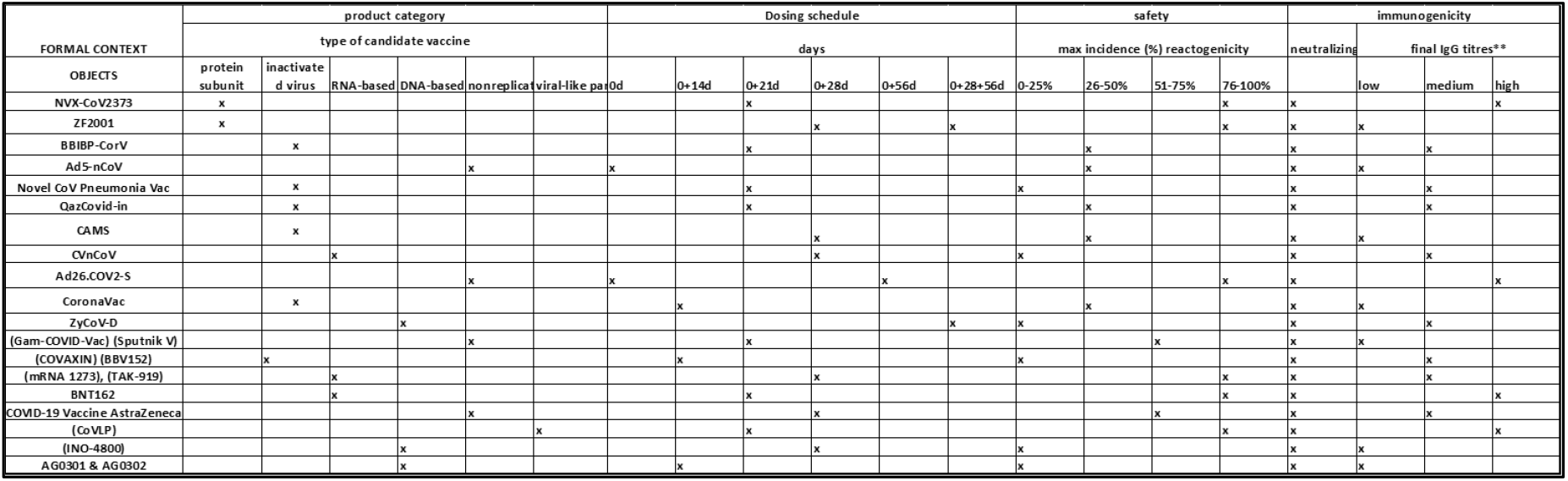
The Formal Multivalued Context for COVID-19 phase 3 Vaccines**-K**: = (V, A, R). For the elaboration of the formal context, 19 vaccine candidates (objects) were used and 4 attributes were defined. To each attribute a nominal scale was assigned, whose values were mutually exclusive. The scales and its values (formulas 2-5) were defined based on the review of the published results of each vaccine.

### 2.2 Formal Concept Analysis

Formal concept analysis (FCA) is a mathematical theory oriented, in particular, at applications in knowledge representation, knowledge acquisition, and data analysis. In [32] R. Wille has introduced Formal Concept Analysis as an application of order and lattice theory in connection with so-called Galois connections induced by relations. The theory is based on a set theoretical model for conceptual hierarchies. This model mathematizes the philosophical understanding of a concept as a unit of thoughts consisting of two parts: the extension and the intension (comprehension). The extension covers all objects (or entities) belonging to the concept, while the intension comprises all attributes (or properties) valid for all the objects under consideration. In the present study the objects (V) correspond to the set of all phase-3 COVID-19 vaccines and their attributes (A) correspond to the technological platform, dosage administration, safety and immunogenicity of each vaccine. Moreover, R (⊆ V × A) is a binary relation between the sets of objects and attributes, respectively. As it is often difficult to list all the objects belonging to a concept and usually impossible to list all its attributes, it is natural to work within a specific context, named formal context, in which the objects and attributes are fixed, here denoted **K**: = (V, A, R). In formal contexts, which usually refer to some application, the relation (v, a) ∈ R (often also written as vRa) is read as follows: The object v is in relation R to the attribute a, if “the object v has the attribute a” for example “the vaccine v is safe”.

A formal context **K** can be considered as (the mathematical model of) a table, which relates objects and attributes of a “real situation”. The entries in the table indicate by a (x) that the object, the name of which precedes the corresponding row, has the attribute, the name of which is at the top of the corresponding column (of the entry). And by an empty space (blanc: “”) it is expressed that the corresponding object does not have that attribute. Once the formal context **K**: = (V, A, R) is constructed, a formal concept (of the context **K)** is derived as follows:

If B⊆ V, D⊆A are arbitrary subsets, then the following derivation operators are defined:

B’= {a∈A/vRa for All v∈B}

D’= {v∈V/vRa for All a∈D}

The pair (B, D) where B⊆V, D⊆A, B’=D and D’=B is called a (formal) concept (of the context **K)** with the extent B and intent D. To obtain the set of all concepts encoded in the formal context, a central result of Formal Concept Analysis [8] is employed, The Fundamental Theorem on Concept Lattices which asserts that given any formal context, it is always possible to obtain a complete lattice that represents it. this complete lattice is called the Concept Lattice of the given context. The proof of the theorem generates an algorithmic procedure that is the basis of the ConExp (java) software [33] to obtain the conceptual Lattice, denoted ⟨**B** (V, A, R);≤⟩, of the defined context, in the present case of **K**: = (V, A, R).

The concept lattice, being a rather universal structure, provides a wealth of information about the relations among objects and attributes, which made possible applications in areas ranging from history and sociology to software engineering and machine learning to e-mail management and ontology building. Indeed, it can help in processing a wide class of data types (for example, any data of phase 3 COVID-19 vaccines represented as a table).

Finally, apart from structural representation of data, the concept lattice provides a framework in which various data analysis and knowledge acquisition techniques can be formulated, in particular through the notion of attribute implication [8]. An implication asserts a certain relationship between two attribute sets, called premise and conclusion: an implication is valid in the data set if every object that has all attributes from the premise of the implication also has all attributes from its conclusion. There is a particular implication cover, called the Duquenne–Guigues basis [34], which is proven to have the minimal size among all covers i.e. The set of all valid implications from which all other implications follow semantically. To carry out the Formal Analysis of Concepts for the COVID-19 vaccines in phase 3 or authorized, the ConExp software (Concept Explorer (Java)) was used, however, today there is an excellent arsenal of free code tools such as Tockit, Galicia, FcaStone, Camelis, Phyton FCA Tool, among others [35].

## 3. Results and discussion

### 3.1 A context for the COVID-19 phase 3 vaccines

To construct the formal context **K**: = (V, A, R), the 19 vaccines listed in table 1 are taken as objects, which then make up the set V and the set of attributes A is defined as follows:

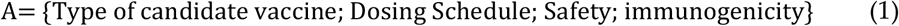

In turn, sub-attributes are defined for each attribute, forming what is called a multivalued context [], for the type of candidate vaccine:

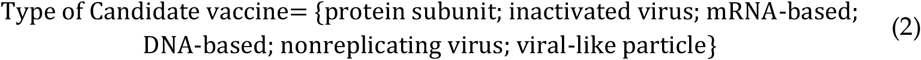

Regarding the dosing Schedule, the assigned values, in days, were the following:

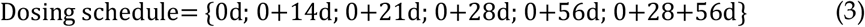

Safety was defined taken into account the maximum percentage (%) of reactogenicity incidence among the vaccinated groups reported by the producers regarding the results of phase1 and phase 2 clinical studies for each of the 19 candidate vaccines.

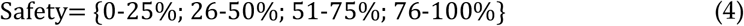

For immunogenicity, the generation of neutralizing IgG antibodies against the complete SARS-CoV-2 virion were considered an essential attribute for all COVID-19 vaccines and also, the final IgG titers, divided in three categories: low, medium and high IgG neutralizing IgG antibodies.

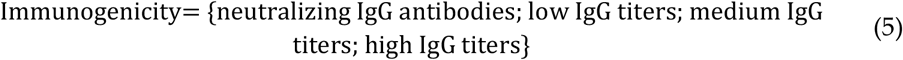

Having defined extension V and intention A for the COVID-19 vaccines in phase 3 and/or authorized, the formal context **K**: = (V, A, R) is elaborated according to the proposed methodology, obtaining the following result:

Reading the context can be done in a simple way, observing that the *ith* vaccine has the *jth* attribute, which is indicated by an *x* in position *ij* of the table. By way of example, it can be seen that the BNT162 vaccine has the attribute of being administered in two doses separated by 21 days apart.

### 3.2 The Vaccine Concept Lattice and its implications

As has been mentioned earlier, a concept for this context consists of an ordered pair (B, D), where B (the *extent*) is a subset of the 19 vaccines and D (the *intent*) is a subset of the four attributes with its sub attributes (formulas 1-5). To demand that the concept is determined by its extent and its intent means that D should contain just those properties shared by all the vaccines in B and similarly, the vaccines in B should be precisely those sharing all the properties in D. A simple procedure for finding a concept is as follows: take an object, say BNT 126, and let D the set of attributes which it possesses, in this case

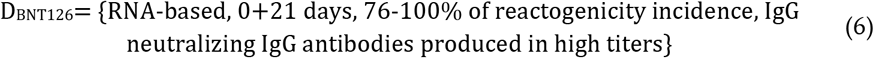

Then let B the set of all vaccines possessing all the attributes in D, in this case

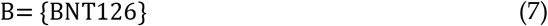

Then (B, D) =({BNT126}, {RNA-based, 0+21 days, 76-100% of reactogenicity incidence, IgG neutralizing IgG antibodies produced in high titers}) is a concept, whose complete lattice representation can be seen in **figure 1**.

**Figure 1.**
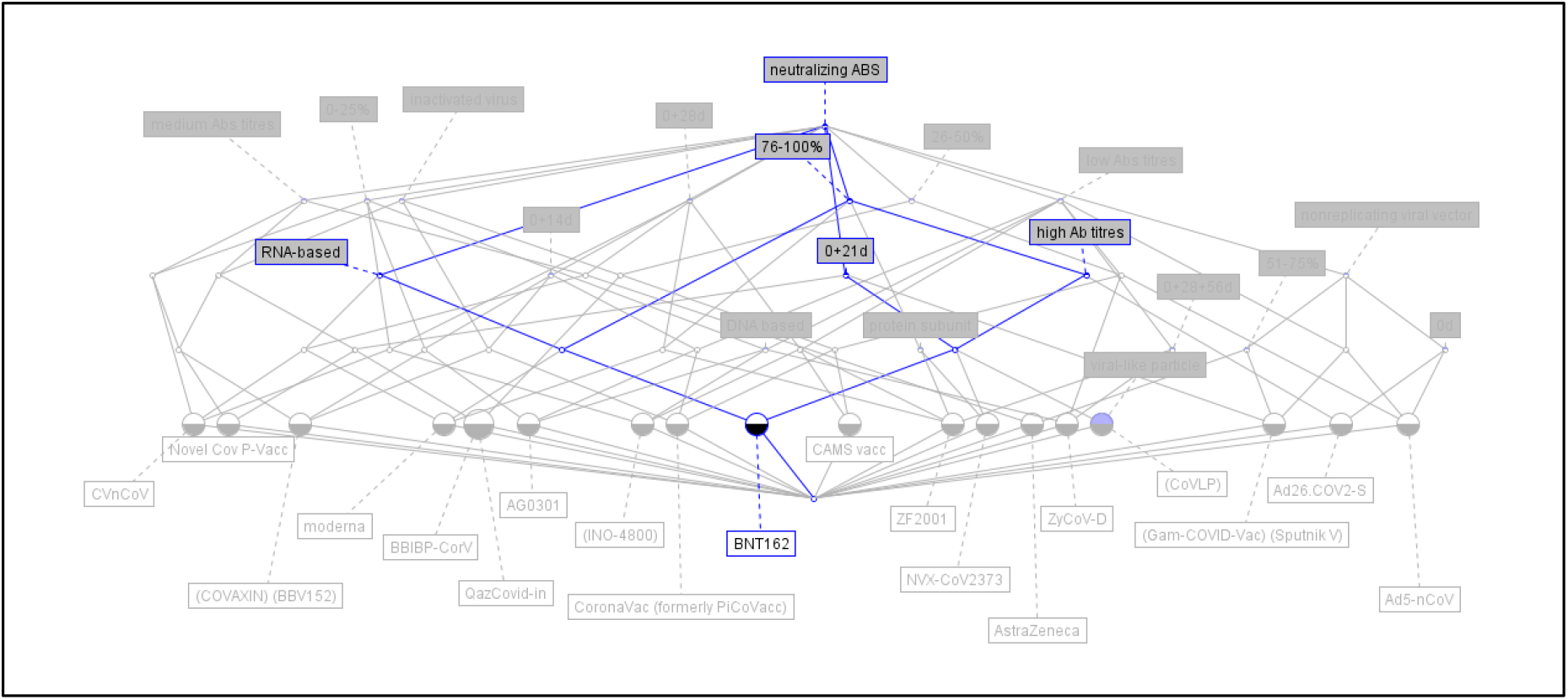
The Concept Lattice of the Pfizer/BioNTec + Fosun Pharma vaccine. The black/white circle represent the concept whose intent can be found by following all line paths going upwards from the circle and noting down the resalted attributes.

On the other hand, employing ConExp, the COVID-19 Vaccine complete Lattice ⟨**B** (V, A, R);≤⟩ (**figure 2)** is obtained from the concept Lattice of the context **K**: = (V, A, R).

**Figure 2.**
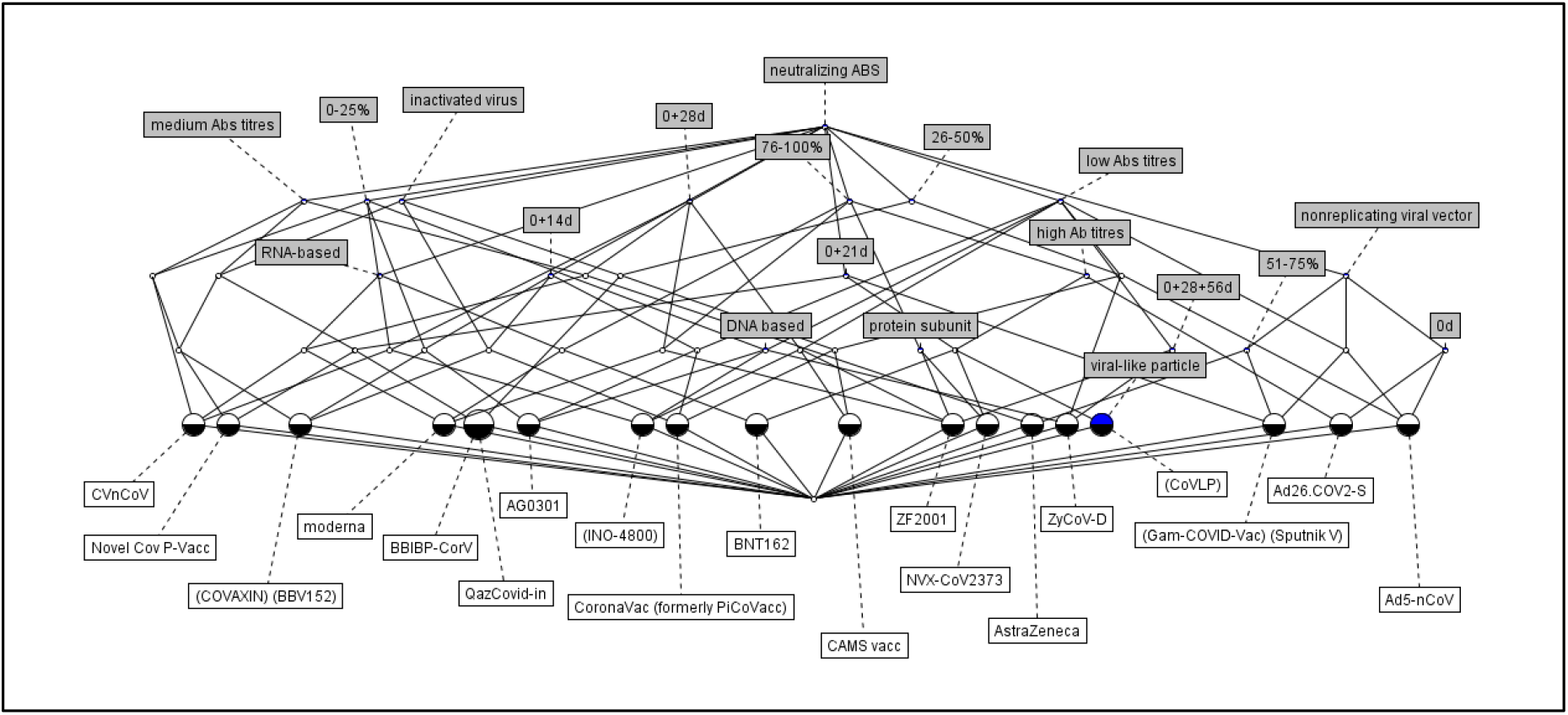
The COVID-19 Vaccine Concept Lattice-⟨**B** (V, A, R);≤⟩ of the context **K**: = (V, A, R).

The COVID-19 Vaccine Concept Lattice-⟨**B** (V, A, R);≤⟩ shows the hierarchical structure that arises from the total of 55 concepts (see supplementary material) that are obtained from the context **K**: = (V, A, R); The Lattice can be read by following the lines that come out of each circle upwards, noting the attributes assigned to each one of them, as already illustrated for the Pfizer BioNTec vaccine. It should be noted that the concept demarcated with the largest diameter circle contains as an extension two objects, the BBIBP-CorV vaccine and the QazCovid-in vaccine; and its intention corresponds to technological platform based on inactivated viruses, with a reactogenicity level of medium incidence (26 −50%), applicable in two doses separated by a period of 21 days, and that generate neutralizing IgG antibodies of medium titers when characterized by ELISA. On the other hand, the Medicago CoVLP vaccine presents a concept marked by a black / blue circle because it is the only vaccine, of those considered in this study, whose technological platform is not shared by any other vaccine.

### 3.3 implications

The power of FCA lies in its ability to build a set of logical implications from the concepts of the context, which constitute the Duquenne-Guiges basis of the COVID-19 Vaccine Concept Lattice-⟨**B** (V, A, R);≤⟩. Given that the implications, derived by ConExp, are written quite schematically, such as: “⟨19⟩ {} ⇒ neutralizing IgG antibodies”, it is necessary to write legibly the set of implications obtained. As an example, and continuing with the previous implication, it is clearer to express it as follows: “All the COVID-19 vaccines in clinical stage 3 generate neutralizing IgG antibodies in vaccinated subjects”. Thus, incorporated language expressions are necessary for the transformation of the potential mathematical to the actual-logic understanding of the conceptual structures [36].

Following this strategy, the implications resulting from the (formal) analysis of concepts are expressed. An important implication is related with reactogenicity, it is observed that the most reactogenic vaccines, i.e., with more than 76% incidence of adverse reactions, are the ones that generate the highest neutralizing antibody titers, they are within this group, Novavax, ZF2001, Janssen’s Ad26.Cov2-S, Moderna’s vaccine, Pfizer BioNTec’s and AstraZeneca’s vaccine. Another implication is that all these vaccines are administered in two doses, mainly in a 0 + 21-day schedule, with the exception of Janssen’s vaccine, which is administered in a single dose.

Taking into account the technological platform on which the vaccines were developed, the following implications can be highlighted:

The six vaccines made from inactivated viruses, namely BBIBP-CorV of Beijing Institute of Biological Products, Novel CoV Pneumonia Vac of Wuhan Institute of Biological Products, QazCovid-in of The Republic of Kazakhstan, CAMS vaccine of the Institute of Medical Biology, Chinese Academy of Medical Sciences, Sinovac, and BBV152 of Bharat Biotech, all are administered in two doses, show less than 50% incidence of adverse reactions among vaccinated patients, and all of them induce neutralizing antibody titers of medium or low levels.

In relation to DNA vaccines, ZyCov-D of Zydus Cadila Healthcare Limited, INO-4800 of Inovio Pharmaceuticals and AG0301 of Osaka University, all presented a low incidence of adverse effects, less than 25%, but also low neutralizing antibody titers against complete virus in follow-up immunoassays in reported clinical studies.

The RNA based vaccines BNT126 from Pfizer / BioNTec and mRNA 1273 from Moderna, induce high antibody titers, high reactogenicity and are applied in the two-dose schedule 0 + 21 days, while the CVnCoV-Curevac vaccine from Bayer is less reactogenic and generates lower antibody titers, but it should be noted that it has an administration scheme of two doses 0 + 28 days, which should be taken into account as a possible explanation for the difference observed; of course it will be necessary to consider the type of adjuvant used that does not it was taken into account in the present study.

Regarding the four vaccines are made from non-replicating viral particles, two of them are applied in a single dose, that of CanSino Biologics (Ad5-nCoV) and that of Janssen (Ad26.CoV2-S), but while the latter induces high reactogenicity (76-100% incidence) and high neutralizing antibody titers, the first shows medium reactogenicity (26-50%) and low neutralizing antibody titers, which suggests an important role for the adjuvants used. On the other hand, Sputnik V of Gamaleya Research Institute and the COVID-19 Vaccine of AstraZeneca, these vaccines are applied in two doses, 0+21 days and 0+28 days respectively, both of them generate 51-75% of adverse reaction incidence among those vaccinated subjects, and show low and medium immunogenicity levels respectively.

The vaccines made from protein subunits, ZF2001 and NVX-Co2327, both generate high reactogenicity (76-100%), however the first, which is applied in two (0 + 28 days) or three doses (0 + 28 + 56 days) induces low levels of neutralizing IgG antibodies, while the second, which is applied under a scheme of 0 + 21 days, induces high titers of neutralizing IgG antibodies against complete viral particles, as evidenced by the results reported for follow-up immunoassays to vaccinations.

Finally, the Medicago vaccine, CoVLP, made from viral-like particles, induces a high reactogenicity (76-100% incidence of adverse reactions), generates high titers of neutralizing IgG antibodies against complete SARS-CoV-2 viral particles and, as has been observed in other vaccines with similar attributes, it is applied under a two-dose scheme, 0 + 21 days.

### 3.4 The dual vaccine concept Lattice

Once the conceptual lattice **B** has been analyzed, which emphasizes the hierarchical structure that exists between concepts, whose objects are vaccines and its attributes are the technological platform, dosage, safety and immunogenicity, resulting in a set of implications on the vaccines given its attributes; it is worth asking what information can be obtained from analyzing the dual case (**B**^**d**^), in which the previous attributes are taken as objects and the previous objects as attributes, seeking to obtain a new set of implications that complement those already obtained. This operation can be easily done with ConExp (**Figure 3**), but its importance lies in the fact of obtaining implications that have biological relevance and not in that it becomes a merely mathematical exercise.

**Figure 3.**
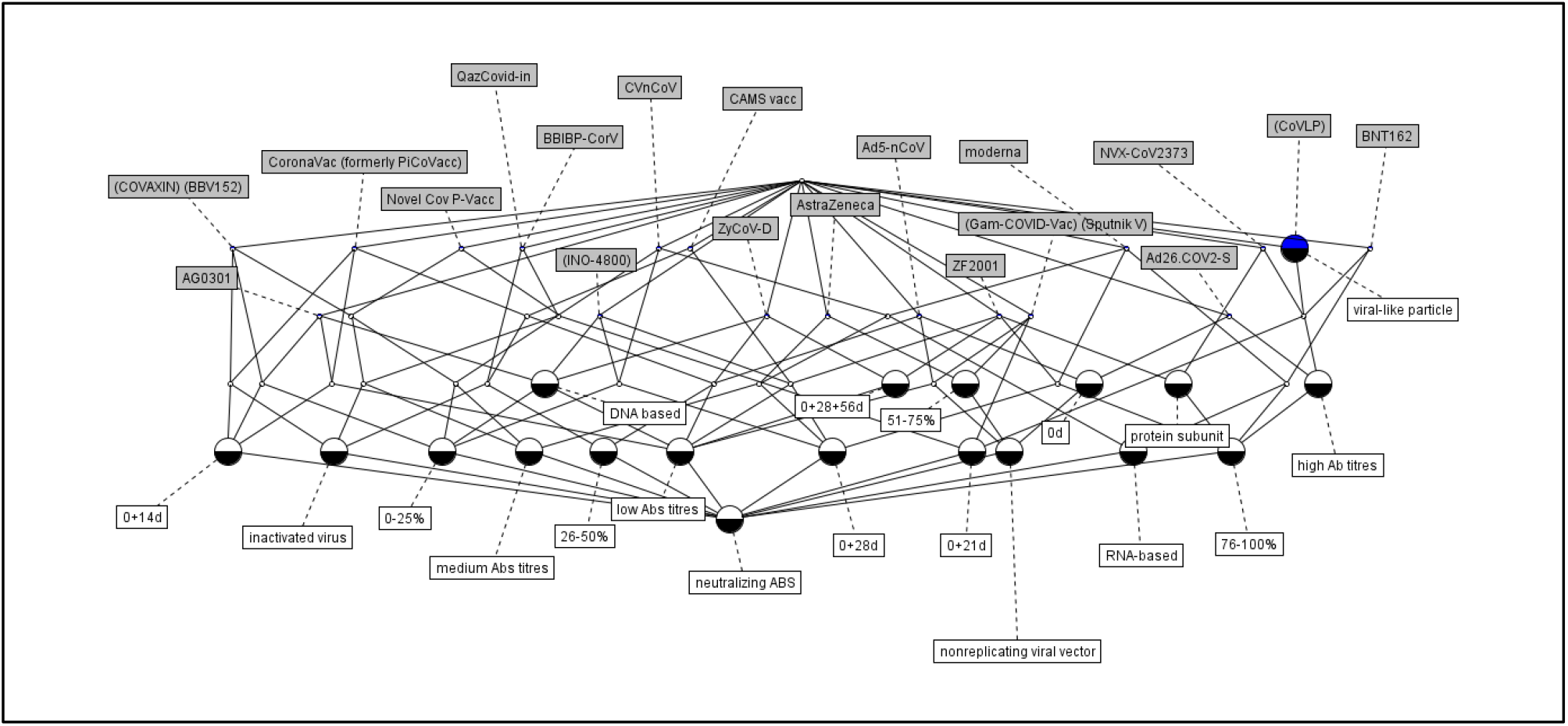
The Dual Vaccine Concept Lattice -⟨**B**^**d**^ (V, A, R);≤⟩ is obtained exchanging the roles of objects and attributes, operation that is always possible to do due to the principle of Duality of the Concept Lattices [8].

In the present work, it is proposed that the biological relevance of the Dual Concept Lattice (**B**^**d**^) lies in the fact that it provides knowledge on how to establish prime-boost immunization strategies. Traditionally, prime-boost immunizations can be given with unmatched vaccine delivery methods while using the same antigen, in a “heterologous” prime-boost format [37]. The most interesting and unexpected finding is that, in many cases, heterologous prime-boost is more effective than the “homologous” prime-boost approach. The pandemics has prompted a very rapid progress of novel vaccination approaches, such as DNA vaccines, RNA vaccines, and viral vector-based vaccines among others, which certainly will expand the scope of heterologous prime-boost vaccination [38], and If some combinations work, they may provide needed flexibility whenever production of a vaccine falters, as often happens. And there’s even a chance that mixing doses of two different vaccines may boost the protection against COVID-19 [39].

#### 3.4.1 Implications for COVID-19 vaccination strategies

From the Dual Vaccine Concept Lattice -⟨**B**^**d**^ (V, A, R);≤⟩ it is possible to obtain 155 logic implications among all the COVID-19 phase 3 vaccines, now viewed as attributes, taking into account their characteristics like safety and immunogenicity, now viewed like objects; Given the hierarchical structure of **B**^**d**^ it is possible to arrange the implications in a decreasing order, in accordance with the bioaffinity of their extension (number of objects shared in common) among the vaccines. The first 40 implications were selected to elaborate a COVID-19 Vaccination Matrix (**Table 3**) given that these include at least 2 or 3 common objects in their extension.

**Table 3.**
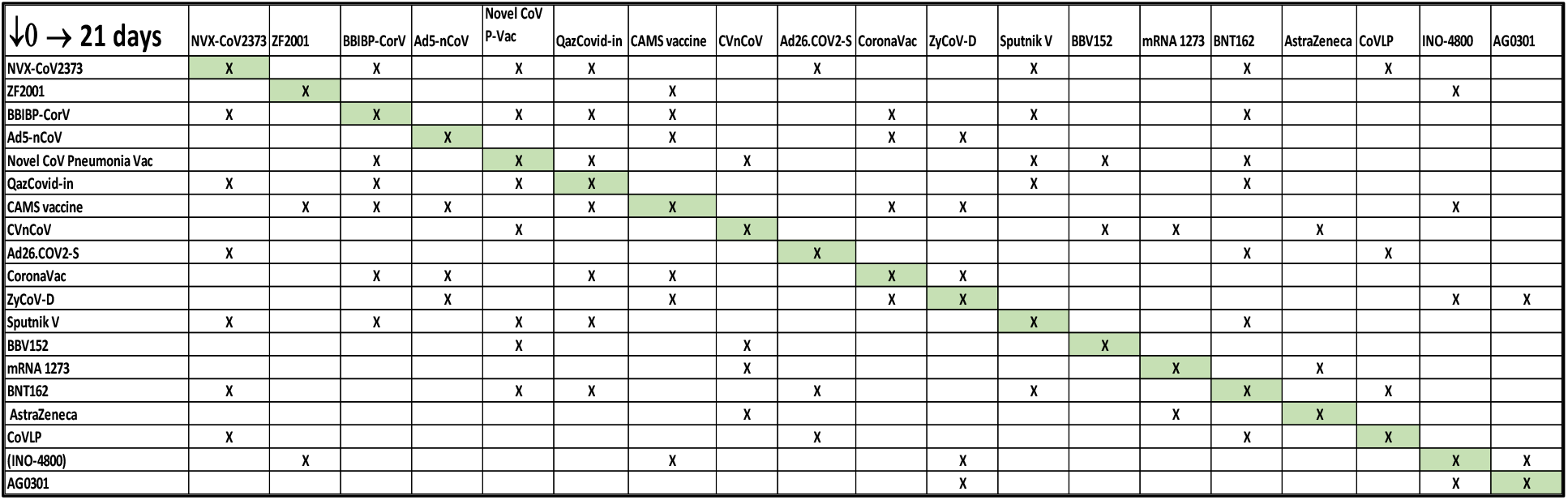
The COVID-19 vaccination matrix is constructed from the analysis of the implications between the different vaccines obtained from the Dual Vaccine Concept Lattice -⟨**B**^**d**^ (V, A, R);≤⟩.

As an example, it is presented here, three of the first 40 implications or association rules, related with Pfizer/BioNTec (BNT162), Medicago (CoVLP) and Novavax (NVX-CoV2373) vaccines,

“⟨4⟩ NVX-CoV2373 BNT162 = [100%] ⇒ ⟨4⟩ CoVLP”

“⟨4⟩ NVX-CoV2373 CoVLP = [100%] ⇒ ⟨4⟩ BNT162”

“⟨4⟩ BNT162 CoVLP = [100%] ⇒ ⟨4⟩ NVX-CoV2373”

Taken together the implications indicate the bioaffinity that relates the three vaccines, which share a total of 4 objects in their extension. It is important to note that the work hypothesis to construct the COVID-19 vaccination matrix is that bioaffine vaccines could be administered in a homologous and also, heterologous schema. To construct the matrix (**Table 3**), proceed as follows: the vaccine is located on the horizontal axis (dose 0) and an “x” is placed under the column containing the vaccine (s) that were related to the first one, which would be administered on the 21st day; in the case of the previous example, the procedure is repeated for each of the three vaccines given its bioaffinity. A total of 98 combinations results from the analysis of the Dual Vaccine Concept Lattice -⟨**B**^**d**^ (V, A, R);≤⟩. As is noted in **Table 3**, the diagonal of the matrix indicates the actual manner the vaccines are administered in a homologous scheme, and all those entries out of the diagonal correspond to possible heterologous vaccination schemes.

It is important to emphasize that this is a very preliminary approach to an answer to the question: Should COVID-19 vaccines be mixed and matched? Of course, a greater number of variables related to dose size, route of application, use of adjuvants, more complete characterization of the specific immune response, both humoral and cellular associated with each vaccine, detailed registry of adverse reactions in each population, should be considered. As vaccine campaigns are carried out worldwide, it will surely be necessary to consider additional variables, here concepts, in order to evaluate the safety and relevance of combining vaccines. Moreover, the safety and effectiveness of mixed vaccines has not be studied and more research is needed, however the methodology proposed here may be useful to determine the form of develop new vaccination schedules to control the pandemic caused by SARS-CoV-2.

## 4. Conclusions

The use of Knowledge Representation, through the Formal Concept Analysis, allows us to reach a series of conclusions about the vaccines for COVID-19 that are in phase 3 or already approved for use. In the first place, this set of vaccines can be represented as a hierarchical structure, in particular as a lattice, The COVID-19 Vaccine Concept Lattice-⟨**B** (V, A, R);≤⟩, based on attributes such as technological platform, dosage, safety and immunogenicity, which is characterized by having a “supremum” that corresponds to the fact that all these vaccines induce neutralizing IgG antibodies against SARS-CoV-2.

The conceptual analysis indicates that there is a direct relationship between the reactogenicity of the vaccines and their ability to induce neutralizing IgG antibodies. The higher the incidence of adverse effects, the higher the antibody titers are obtained. of course, reactogenicity generally consists of minor discomforts such as redness, fever, pain and others widely described elsewhere in the literature []. It should also be noted that the most effective dosage, regardless of the technological platform of the vaccine, is that of two doses delivered 21 days apart.

Regarding the technological platform of COVID-19 vaccines in stage 3 of clinical evaluation, it is observed that vaccines made from inactivated viruses show an incidence of adverse reactions of less than 50% in vaccinated individuals and generate medium or low titers of neutralizing IgG antibodies. DNA vaccines show even less reactogenicity, all less than 25% incidence, and induce low levels of neutralizing IgG antibodies. In contrast, RNA vaccines are strongly reactogenic and tend to induce high titers of neutralizing IgG antibodies.

As for non-replicating viral particle vaccines, their reactogenicity is medium to high level (50% or more incidence), and they consistently induce, in general, medium to high neutralizing antibody titers. However, it is important to note that this type of vaccine seems to be more effective when administered in two doses under the 0 + 21-day schedule. The same type of behavior is observed for the protein subunit vaccines, such as ZF2001 and NOVAVAX, as well as for the CoVLP vaccine made from viral-like particles technology.

The Dual Vaccine Concept Lattice -⟨**B**^**d**^ (V, A, R);≤⟩ allows to establish the level of affinity within the set of vaccines, making possible the development of a methodology for the elaboration of heterologous vaccination strategies for the control of the pandemic, based on the previous result that relates a better neutralizing humoral response with the use of two doses applied 0 + 21 days. The vaccination strategy is consolidated through what is called the COVID-19 vaccination matrix/space based on the logical implications of the dual concept lattice.

## Data Availability

there are no new data associated with this paper

